# Contributing factors associated with violence against children and intimate partner violence against women in households in southern Senegal

**DOI:** 10.64898/2026.01.15.26344174

**Authors:** Bryan Shaw, Erin DeGraw, Christina Yantis, Fatou Yate Gueye, Mor Lo, M. Badiene, Jamie Greenberg, Anjalee Kohli

## Abstract

**Background:** Intimate partner violence (IPV) and violent discipline of children frequently co-occur, but are rarely addressed jointly in public health research or programming. In Senegal, both IPV and violent discipline are prevalent, particularly in rural regions, yet little is known about their intersection within households. This study investigates factors associated with IPV and violent discipline among young couples in southeastern Senegal and explores pathways between violence-related attitudes, social norms, IPV, and violence against children.

**Methods:** We conducted a cross-sectional baseline survey of 330 heterosexual couples (fathers aged 18–35 and their partners) in two rural communes in Senegal as part of a quasi-experimental evaluation of the REAL Fathers Initiative. Structured interviews assessed self-reported IPV, violent discipline of under-five children, social norms, parenting practices, and relationship quality. Multivariate logistic regressions and structural equation modeling (SEM) were used to identify predictors of IPV and violent discipline and test hypothesized pathways between them.

**Results:** Over half of couples (57%) reported IPV in the past three months; 89% of mothers and 71% of fathers reported using violent discipline. Mothers reported more frequent use of physical and harsh discipline than fathers. Among men, attitudes and injunctive norms supporting violence were significantly associated with physical discipline. Among women, higher education and confidence in nonviolent parenting were associated with lower likelihood of violent discipline. SEM showed a significant pathway from IPV to fathers’ use of physical violent discipline, with additional indirect effects through maternal violence.

**Conclusions:** Findings reveal high co-occurrence of IPV and violent discipline, shared risk factors, and evidence of interlinked pathways between partner and child-directed violence. These results underscore the need for integrated interventions targeting both forms of violence within the family system and support social norm change and parenting skills among young couples.

## Introduction

Globally, an estimated 27% of ever-partnered women aged 15-49 years have experienced physical and/or sexual intimate partner violence (IPV) by a male partner [1]. Peterman et al. [2] analyzed nationally representative IPV data from 30 countries and reported that abuse starts on average, 3.5 years into union among ever-married women who experience abuse after union. For many women, IPV may start in the first pregnancy or early in the pregnancy, indicating the risk of experiencing IPV for younger women [3].

Women in Sub-Saharan Africa report the largest global burden of lifetime experience of physical and/or sexual IPV, between 27-44% [1]. An analysis of Demographic and Health Surveys (DHS) in Africa reported prevalence of IPV was associated with being 25-34 years while other potential risk factors for IPV included women having lower levels of education, employment (for women), religion, living in a rural area, poverty, having more children, traditional gender role beliefs, and less engagement in decision making [4]. The 2019 Senegal DHS reported 16.9% of women aged 15-49 years ever experienced any IPV. Rural-urban disparities in IPV prevalence vary by region, with higher prevalence in the South (26.6%) and Central (18.7%) regions [5]. One-third of women reporting IPV in Dakar had one child and one-quarter had two children, indicating that IPV starts early in the relationship [6]. A recent study documented increased individual acceptance of IPV among those with social networks that considered IPV acceptable [7]. This suggests that interventions seeking to prevent IPV in Senegal may need to consider the social network and their influence on violence-related attitudes, prevention programming, and help seeking.

### Violent discipline

Between 133 and 275 million children frequently witness violence between their caregivers [8]. Growing up in a home with IPV increases risk for child maltreatment [9–11]. An analysis of 21 United Nations Children’s Fund (UNICEF) Multiple Indicator Cluster Survey (MICS) surveys reported that in 41% of households where men and women agree that hitting or beating a wife is justified, they also consider corporal punishment as a necessary part of child rearing [12]. Of children between 2 and 14 years globally, about 6 in 10 regularly experience physical punishment by their caregivers [13]. In Senegal, a household survey in two departments in Senegal (Pikine, Kolda) found that 34.5% and 26.8% of adolescents experienced an adult in the home being physically and emotionally, respectively, violent towards a child [14]. A five-country study conducted by the African Child Policy Forum reported that between 66% to 90% of girls experienced physical violence by anyone before the age of 18 years. Girls also reported psychological violence before age 18, most frequently perpetrated by their parents, siblings, peers, and teachers [15].

Research data show that parents use multiple strategies to discipline their children, violent and non-violent discipline. Possible reasons for use of violent discipline vary, including social norms on the acceptability or expected use of violent discipline as a strategy for child rearing [16], expectations of child development and behavior relative to their age, and lack of knowledge of alternate forms of discipline [17, 18]. Violence at young ages is associated with increased risk for physical injury, children’s inability to understand the action or the motivation behind it, or to adopt coping strategies [19]. Studies from high-income countries indicate long-term outcomes associated with physical punishment. These include externalizing behaviors, aggressive behaviors, conduct problems, depressive symptoms, poorer cognitive outcomes, and lower quality parent-child interaction [20]. Along with a range of health and well-being outcomes, children who experience violence themselves or grow up in a home with IPV are at increased risk of experiencing or using violence in their future relationships [11, 21].

### Preventing the co-occurrence of violence

Despite their frequent co-occurrence [21], IPV and corporal punishment have often been addressed separately. REAL Fathers is one intervention designed specifically to engage with men as partners and fathers, seeking to prevent IPV and violent discipline of young children. REAL Fathers engages young fathers who are parenting young children. The program is designed to reach men before social norms related to gender roles, attitudes, expectations, and behaviors in parenting and partnership are established. Young fathers identify a respected elder male to be trained as their mentor; mentors engage fathers in one-on-one, couples, and group mentorship to build knowledge, skills, peer support, and behavior change. The program is supported by an emotion-based poster campaign and a public celebration event.

REAL Fathers was evaluated twice in a quasi-experimental study with men [22] and in a randomized control trial with men in Uganda [23]. Both studies demonstrated the effectiveness of REAL Fathers to improve couple communication, prevent IPV, increase fathers’ caregiving role, and prevent violent discipline. The adaptation and evaluation of the REAL Fathers Initiative (2020-2022) in Senegal and Rwanda was supported by the United States Agency for International Development (USAID)-funded Partnerships Plus program, through John Snow International, to Plan International and the Institute of Reproductive Health at Georgetown University. Key adaptations to REAL Fathers for Senegal included expanding eligibility of young fathers 18-35 years with under-five children and adjustments for cultural context and language. In addition, to address evidence gaps on IPV prevention from the evaluation in Uganda, this study includes both men participating in REAL and their female partners.

Using baseline data from the REAL Fathers, this study deepens the understanding of household violence in Senegal. Specifically, this study includes three objectives. First, to understand the individual and relationship factors associated with the father’s and mother’s use of violent discipline with their under-five child(ren). Second, to understand the individual and relationship factors associated with men’s perpetration and women’s experience of IPV. Third, to understand the potential pathways between attitudes, norms, relationship factors, IPV, and mother’s and father’s use of violent discipline.

## Materials and Methods

### Study design

This baseline survey is part of a quantitative, quasi-experimental pre-/post-test evaluation of REAL Fathers in two Communes in southeastern Senegal: Kedougou and Tomboronkoto. In each Commune, eight villages were selected based on accessibility, relationships between Plan-Senegal, the local government, and organizations, and community interest in participating in REAL Fathers.

Recruitment of young fathers to participate in REAL took place before the baseline survey from May 20 to August 15, 2021. The baseline survey took place in November 2021 and included interviews with couples; men and women interviewed separately.

### Eligibility criteria

Eligibility criteria were determined by REAL’s focus on young men newer to marriage and parenting. Young fathers were eligible for intervention and study if they were: residents of a selected Commune and village; aged 18-35 years; married or living with a female partner 18 years or older; had at least one child 0-5 years of age; and committed to participating in REAL. Fathers were identified through consultation with local community leaders and Plan-Senegal’s networks. Female partners of selected fathers were eligible for participation in the baseline survey. Interviews were conducted in French or Pulaar in person by trained interviewers conducting interviews with participants of the same gender. *Ethical considerations.* The study was reviewed and approved by the Georgetown University Institutional Review Board (STUDY 00003454) and the Comité National d’Éthique pour la Recherche en Santé (CNERS) of Senegal (STUDY SEN18/61). All participants gave voluntary, written informed consent, and interviews were conducted in a private location. ID codes were used in place of participant names. Interviewers were trained in sensitive and ethical interviewing, including data security and safety associated with interviewing a couple.

### Study measures

Mothers and fathers self-reported their violent discipline of children 0-5 years in the past three months. Measures were adapted from the Parent-Child Conflict Tactics Scale [24], a tool that is incorporated into the UNICEF MICS. Psychological aggression against children 0-5 years included four items, physical punishment included six items, with two of these items taken to signify harsh physical punishment. All items were assessed on a 4-point scale (never or rarely, once in a while, several times a week, every day). We considered perpetrating any act of violent discipline against under-five children, at any frequency more than never/rarely, in the previous three months as a measure of each type of violent discipline.

Men’s reported perpetration and women’s reported experience of physical (6 items), sexual (3 items), and psychological (4 items) IPV and coercive behaviors (7 items) in the previous three months were taken from the World Health Organization (WHO) Multi-Country Study on Women’s Health and Domestic Violence Against Women Study [25]. All items were assessed on a 4-point Likert scale. Reports of experiencing (women) or perpetrating (men) any act of emotional, physical, or sexual IPV in the previous three months, of any frequency more than never/rarely, was considered an indication of each type of IPV. We also considered IPV within the couple dyad, and a household would be considered to include IPV if either the man reported perpetrating IPV and/or the woman reported experiencing IPV in the previous three months. For both violent discipline and IPV, a period of three months of recall was used to align the reporting with the endline assessment, which would be conducted immediately after the project.

Attitudes towards violent discipline towards young children and IPV were drawn from the DHS with a 4-point Likert response [5]. Descriptive norms were measured with two items each, with 4-point Likert response options relating to perceptions of typical behaviors in respondent communities about using VAC or IPV. Injunctive norms were measured with four items each, with 4-point Likert response options assessing respondent perceptions about expectations or acceptability of important social others regarding VAC or IPV. Participants were asked about their own confidence in handling child(ren) less than five years of age without resorting to violence (one item, 4-point Likert response), father’s use of nurturing care (six items, 4-point Likert responses), and parent’s use of positive reinforcement (five items, 4-point Likert responses). Additional covariates assessed include age, education, polygamous marriage, total number of children in partnership, household food security, reports of men’s alcohol consumption, and self-rated relationship quality (4-point Likert response) over the last one month.

### Analysis

All data analyses were performed using Stata 16 (StataCorp. 2019. College Station, TX: StataCorp LLC.). Descriptive analysis and bivariate and multivariate logistic regressions were run for all dependent and independent variables and covariates. Dependent variables, including any reporting of VAC or any reporting of perpetration (if male respondent) or experience (if female respondent) of IPV, were treated as binary outcome variables. Any reports by either men, women, or both of IPV in a couple dyad were used as the outcome of interest for both predictors of men’s behaviors (using men’s survey data) and women’s behaviors. This was considered the most reliable indicator of IPV, as women are more likely to report IPV experience than men are to report perpetrating IPV. Multivariate logistic regression models controlled for other independent predictors for VAC and IPV and sociodemographic covariates, and were run separately for men and women.

Structural equation modeling (SEM) was used to develop two models: one for men’s use of physical violent discipline and the other for women’s use of physical violent discipline on their children, five or less years of age. However, only the men’s model had acceptable fit statistics and is presented in this paper. This model used a more comprehensive IPV report, where any household where women reported experiencing any IPV or men reported perpetrating any IPV was considered as having IPV in the relationship. The men’s model assessed the pathways between any IPV reported by either couple member and the father’s or mother’s reported perpetration of physical VAC in the previous three months. Using Stata 16, the robust maximum likelihood method was used to correct the standard errors for some non-normality in the data. The fit indices root-mean-squared error of approximation (RMSEA), standardized root mean squared residual (SRMR), comparative fit index (CFI), Tucker–Lewis Index (TLI), and chi-squared indices were used to assess fit of each model with the data.

## Results

### Socio-demographic data

The baseline study included 330 couples (Table 1). On average, men were 28.8 years and women 21.8 years old. About one-third of the couples only had one child, 8.5% of couples were part of polygamous marriages, and nearly all were practicing Muslims. Over 40% of couple members reported food insecurity at least once per week. Alcohol consumption was rarely reported. Over three-quarters (75.5%) of men and 20.3% of women reported that their relationship was very good.

**Table 1.**
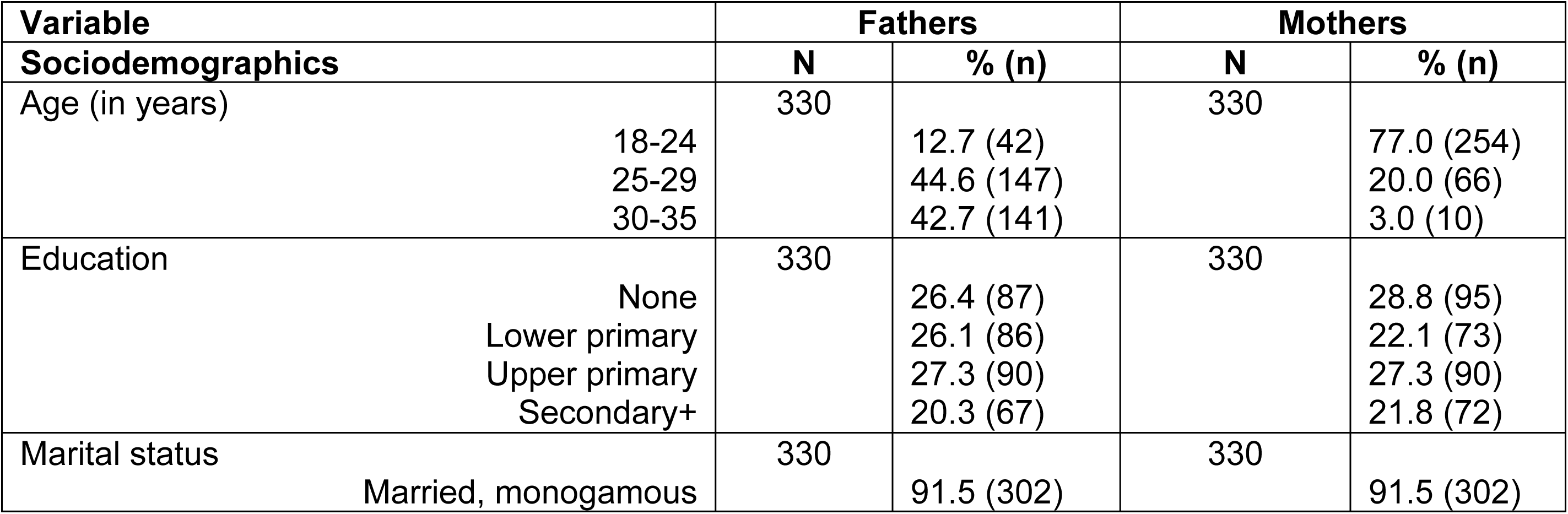
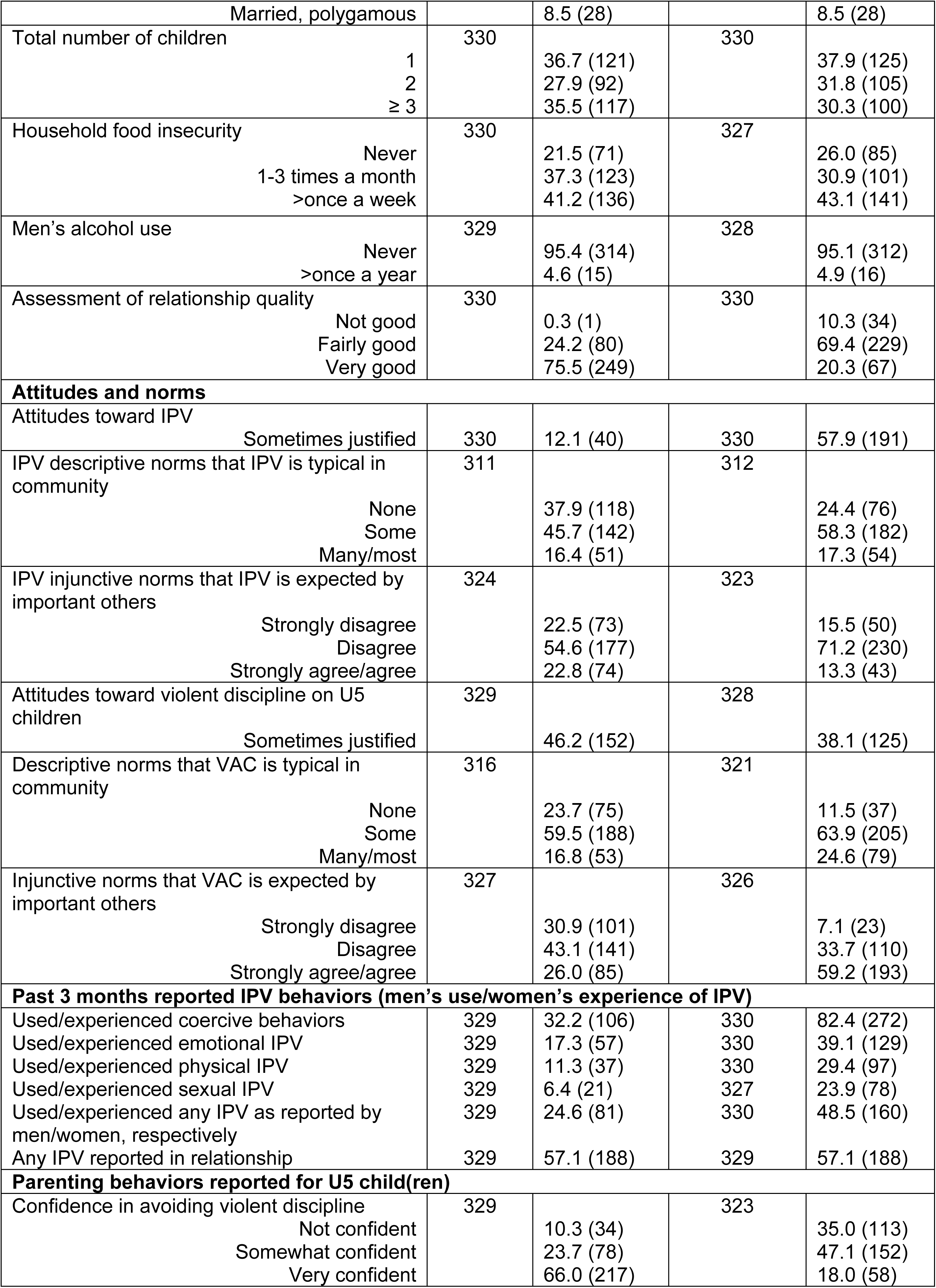
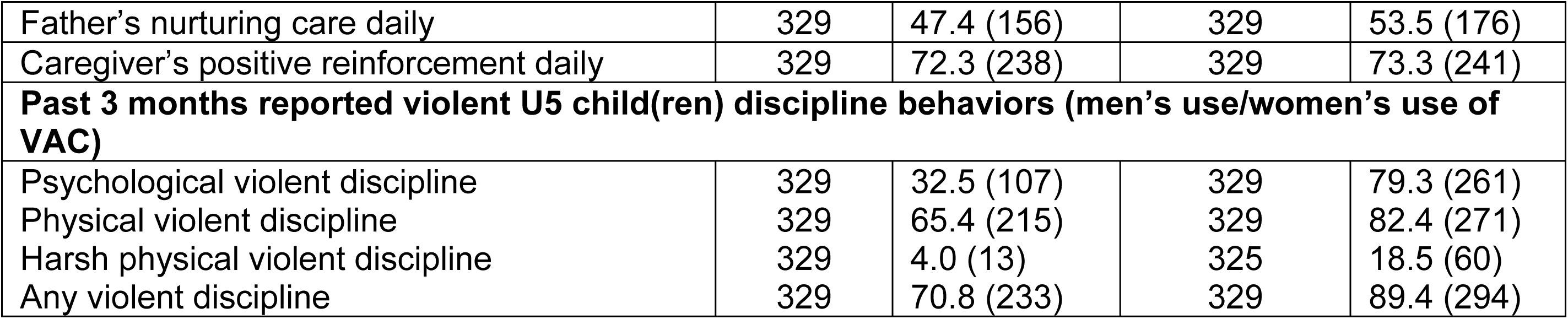
Socio-demographic and behavioral descriptive statistics for fathers and mothers at baseline

| Variable |  | Fathers |  | Mothers |  |
| --- | --- | --- | --- | --- | --- |
| Sociodemographics |  | N | % (n) | N | % (n) |
| Age (in years) | 18-24 | 330 | 12.7 (42) | 330 | 77.0 (254) |
|  | 25-29 |  | 44.6 (147) |  | 20.0 (66) |
|  | 30-35 |  | 42.7 (141) |  | 3.0 (10) |
| Education | None | 330 | 26.4 (87) | 330 | 28.8 (95) |
|  | Lower primary |  | 26.1 (86) |  | 22.1 (73) |
|  | Upper primary |  | 27.3 (90) |  | 27.3 (90) |
|  | Secondary+ |  | 20.3 (67) |  | 21.8 (72) |
| Marital status |  | 330 |  | 330 |  |
| Married, monogamous |  |  | 91.5 (302) |  | 91.5 (302) |

|  |  |  |  |  |
| --- | --- | --- | --- | --- |
| Married, polygamous |  | 8.5 (28) |  | 8.5 (28) |
| Total number of children | 330 |  | 330 |  |
| 1 |  | 36.7 (121) |  | 37.9 (125) |
| 2 |  | 27.9 (92) |  | 31.8 (105) |
| ≥ 3 |  | 35.5 (117) |  | 30.3 (100) |
| Household food insecurity | 330 |  | 327 |  |
| Never |  | 21.5 (71) |  | 26.0 (85) |
| 1-3 times a month |  | 37.3 (123) |  | 30.9 (101) |
| >once a week |  | 41.2 (136) |  | 43.1 (141) |
| Men's alcohol use | 329 |  | 328 |  |
| Never |  | 95.4 (314) |  | 95.1 (312) |
| >once a year |  | 4.6 (15) |  | 4.9 (16) |
| Assessment of relationship quality | 330 |  | 330 |  |
| Not good |  | 0.3 (1) |  | 10.3 (34) |
| Fairly good |  | 24.2 (80) |  | 69.4 (229) |
| Very good |  | 75.5 (249) |  | 20.3 (67) |
| **Attitudes and norms** |  |  |  |  |
| Attitudes toward IPV |  |  |  |  |
| Sometimes justified | 330 | 12.1 (40) | 330 | 57.9 (191) |
| IPV descriptive norms that IPV is typical in community | 311 |  | 312 |  |
| None |  | 37.9 (118) |  | 24.4 (76) |
| Some |  | 45.7 (142) |  | 58.3 (182) |
| Many/most |  | 16.4 (51) |  | 17.3 (54) |
| IPV injunctive norms that IPV is expected by important others | 324 |  | 323 |  |
| Strongly disagree |  | 22.5 (73) |  | 15.5 (50) |
| Disagree |  | 54.6 (177) |  | 71.2 (230) |
| Strongly agree/agree |  | 22.8 (74) |  | 13.3 (43) |
| Attitudes toward violent discipline on U5 children | 329 |  | 328 |  |
| Sometimes justified |  | 46.2 (152) |  | 38.1 (125) |
| Descriptive norms that VAC is typical in community | 316 |  | 321 |  |
| None |  | 23.7 (75) |  | 11.5 (37) |
| Some |  | 59.5 (188) |  | 63.9 (205) |
| Many/most |  | 16.8 (53) |  | 24.6 (79) |
| Injunctive norms that VAC is expected by important others | 327 |  | 326 |  |
| Strongly disagree |  | 30.9 (101) |  | 7.1 (23) |
| Disagree |  | 43.1 (141) |  | 33.7 (110) |
| Strongly agree/agree |  | 26.0 (85) |  | 59.2 (193) |
| **Past 3 months reported IPV behaviors (men's use/women's experience of IPV)** |  |  |  |  |
| Used/experienced coercive behaviors | 329 | 32.2 (106) | 330 | 82.4 (272) |
| Used/experienced emotional IPV | 329 | 17.3 (57) | 330 | 39.1 (129) |
| Used/experienced physical IPV | 329 | 11.3 (37) | 330 | 29.4 (97) |
| Used/experienced sexual IPV | 329 | 6.4 (21) | 327 | 23.9 (78) |
| Used/experienced any IPV as reported by men/women, respectively | 329 | 24.6 (81) | 330 | 48.5 (160) |
| Any IPV reported in relationship | 329 | 57.1 (188) | 329 | 57.1 (188) |
| **Parenting behaviors reported for U5 child(ren)** |  |  |  |  |
| Confidence in avoiding violent discipline | 329 |  | 323 |  |
| Not confident |  | 10.3 (34) |  | 35.0 (113) |
| Somewhat confident |  | 23.7 (78) |  | 47.1 (152) |
| Very confident |  | 66.0 (217) |  | 18.0 (58) |
| Father's nurturing care daily | 329 | 47.4 (156) | 329 | 53.5 (176) |
| Caregiver's positive reinforcement daily | 329 | 72.3 (238) | 329 | 73.3 (241) |
| **Past 3 months reported violent U5 child(ren) discipline behaviors (men's use/women's use of VAC)** |  |  |  |  |
| Psychological violent discipline | 329 | 32.5 (107) | 329 | 79.3 (261) |
| Physical violent discipline | 329 | 65.4 (215) | 329 | 82.4 (271) |
| Harsh physical violent discipline | 329 | 4.0 (13) | 325 | 18.5 (60) |
| Any violent discipline | 329 | 70.8 (233) | 329 | 89.4 (294) |

### IPV and VAC attitudes and norms

Attitudes considering IPV as sometimes justified within a relationship were more common among women (57.9%) compared to men (12.1%). About two-thirds of men (62.1%) and over three-quarters of women (75.6%) reported that IPV is common in some couples in their community, with about 15% reporting that IPV occurs in many or most couples in their community. For injunctive norms, almost one-quarter (22.8%) of men and 13.3% of women agreed or strongly agreed that IPV is acceptable to people in their community. Attitudes favoring violent discipline on children were common, with 46.2% of men and 38.1% of women believing that VAC is sometimes justified for under-five children. Injunctive norms that violent discipline is expected were common among women (59.2%) compared to men (26.0%). Women were more likely to report that many or most parents in their community practice violent discipline against under-five children (24.6%) compared to their male partners (16.8%).

### IPV behaviors

Women reported experiencing physical (29.4%), psychological (39.1%), and sexual (23.9%) IPV in the past three months at least twice as often as men reported perpetrating these behaviors (11.3%, 17.3%, 6.4%, respectively). In 57.1% of the couple pairings, either the male or female couple member reported IPV within the relationship. Coercive behaviors perpetrated by male partners, which are often associated with IPV, were very commonly reported by women (82.4%) while less commonly reported by men themselves (32.2%).

### VAC and parenting behaviors

Whereas fathers were very confident in avoiding violent discipline when correcting their young children, mothers were less likely to report being very confident (18.0%). Nearly three-quarters of fathers (72.3%) and mothers (73.3%) reported an act of positive reinforcement, themselves, daily. Women reported using violent discipline with young children more commonly than men, including psychological aggression (79.3% vs. 32.5%), physical discipline (82.4% vs. 65.4%), and harsh physical discipline (18.5% vs. 4.0%), respectively. Overall, 89.4% of women and 70.8% of men reported using some form of violence to discipline their under-five children.

### Bivariate and multivariate analysis for fathers’ and mothers’ use of physical discipline against under-five children

Table 2 and Table 3 include bivariate and multi-level modeling for men’s and women’s reported use of any violent discipline with their under-five. Men were more likely to report physical VAC if they were not confident in avoiding or personally approved of violent discipline and perceived VAC to be both typical in their community and expected from them in their household in bivariate models. In multivariate models, only personal attitudes approving of VAC (adjusted odds ratio (aOR)=2.18; p<0.01) and perceiving VAC to be acceptable (aOR=1.62; p<0.05) remained significantly associated with higher likelihood of physical VAC among fathers.

**Table 2:** Bivariate and multivariate logistic regression models for men’s behaviors: men’s use of violent discipline against under-five children and perpetration of any IPV against their partner in the past three months

|  | Any physical VAC in past 3 months |  |  |  | Any IPV reported in relationship in past 3 months |  |  |  |
| --- | --- | --- | --- | --- | --- | --- | --- | --- |
|  | OR (95% CI) | P-val. | AOR (95% CI)<sup>a</sup> | P-val. | OR (95% CI) | P-val. | AOR (95% CI)<sup>b</sup> | P-val. |
| **Sociodemographics** |  |  |  |  |  |  |  |  |
| Age | 0.88 (0.62, 1.25) | 0.465 | 0.89 (0.59, 1.35) | 0.594 | 0.93 (0.64, 1.39) | 0.690 | 0.95 (0.61, 1.48) | 0.833 |
| Education | 1.07 (0.86, 1.33) | 0.556 | 1.08 (0.83, 1.40) | 0.561 | 1.05 (0.83, 1.32) | 0.679 | 1.08 (0.81, 1.44) | 0.616 |
| In polygamous relationship | 2.61 (0.97, 7.07) | 0.059 | 2.65 (0.89, 7.85) | 0.079 | 2.97 (1.35, 6.55) | **0.007** | 2.76 (1.03, 7.43) | **0.044** |
| Total number of children | 1.31 (0.99, 1.74) | 0.062 | 1.32 (0.95, 1.83) | 0.103 | 1.68 (1.23, 2.29) | **0.001** | 1.53 (1.06, 2.21) | **0.022** |
| Household food insecurity | 1.17 (0.86, 1.59) | 0.322 | 1.31 (0.89, 1.92) | 0.174 | 0.76 (0.55, 1.06) | 0.105 | 0.93 (0.62, 1.39) | 0.717 |
| Relationship quality | 0.86 (0.50, 1.49) | 0.595 | 1.01 (0.54, 1.87) | 0.978 | 0.28 (0.16, 0.47) | **0.000** | 0.38 (0.21, 0.70) | **0.002** |
| **VAC related factors** |  |  |  |  |  |  |  |  |
| Father's nurturing care | 2.00 (1.23, 3.27) | **0.006** | 1.57 (0.89, 2.76) | 0.120 | - | - | - | - |
| Caregiver's positive reinforcement | 1.58 (0.94, 2.65) | 0.082 | 1.65 (0.92, 2.97) | 0.091 | - | - | - | - |
| Confidence in avoiding violent discipline toward U5 child | 0.66 (0.45, 0.98) | **0.039** | 0.87 (0.51, 1.47) | 0.597 | - | - | - | - |
| Attitudes toward violent discipline on U5 children | 2.26 (1.39, 3.67) | **0.001** | 2.18 (1.29, 3.71) | **0.004** | - | - | - | - |
| VAC descriptive norms | 2.01 (1.34, 3.02) | **0.001** | 1.34 (0.83, 2.18) | 0.229 | - | - | - | - |
| VAC injunctive norms | 1.84 (1.31, 2.57) | **0.000** | 1.62 (1.01, 2.59) | **0.045** | - | - | - | - |
| **IPV related factors** |  |  |  |  |  |  |  |  |
| Coercive behavior toward wife | - | - | - | - | 5.53 (3.23, 9.45) | **0.000** | 3.00 (1.55, 5.81) | **0.001** |
| Attitudes toward IPV | - | - | - | - | 6.11 (3.04, 12.24) | **0.000** | 3.67 (1.65, 8.19) | **0.001** |
| IPV descriptive norms | - | - | - | - | 1.66 (1.15, 2.39) | **0.006** | 0.91 (0.54, 1.54) | 0.730 |
| IPV injunctive norms | - | - | - | - | 2.05 (1.38, 3.05) | **0.000** | 1.13 (0.65, 1.97) | 0.657 |
| <sup>a</sup> Models adjusted for age, education, HH food insecurity, total children in HH & VAC covariates in table |  |  |  |  |  |  |  |  |
| <sup>b</sup> Models adjusted for age, education, HH food insecurity, total children in HH & IPV covariates in table |  |  |  |  |  |  |  |  |

**Table 3:** Bivariate and multivariate logistic regression models for women’s behaviors: women’s use of violent discipline against under-five children and experience of any IPV against from their partner in the past three months

|  | Any physical VAC in past 3 months |  |  |  | Any IPV reported in relationship in past 3 months |  |  |  |
| --- | --- | --- | --- | --- | --- | --- | --- | --- |
|  | OR (95% CI) | P-val. | aOR (95% CI)<sup>a</sup> | P-val. | OR (95% CI) | P-val. | aOR (95% CI)<sup>b</sup> | P-val. |
| **Sociodemographics** |  |  |  |  |  |  |  |  |
| Age | 0.72 (0.39, 1.36) | 0.314 | 0.57 (0.26, 1.28) | 0.176 | 0.62 (0.40, 0.97) | **0.036** | 0.82 (0.46, 1.47) | 0.509 |
| Education | 0.56 (0.40, 0.80) | **0.001** | 0.47 (0.29, 0.76) | **0.002** | 0.84 (0.69, 1.02) | 0.071 | 1.03 (0.81, 1.31) | 0.821 |
| In polygamous relationship | 2.97 (0.69, 12.89) | 0.146 | 1.81 (0.34, 9.68) | 0.486 | 3.52 (1.45, 68.52) | **0.005** | 2.86 (1.00, 8.21) | 0.050 |
| Total number of children | 2.62 (1.54, 4.45) | **0.000** | 4.41 (2.14, 9.06) | **0.000** | 1.12 (0.86, 1.45) | 0.412 | 1.01 (0.72, 1.42) | 0.967 |
| Household food insecurity | 0.58 (0.35, 0.94) | **0.028** | 0.44 (0.24, 0.83) | **0.011** | 1.40 (1.07, 1.84) | **0.014** | 1.27 (0.91, 1.79) | 0.160 |
| Relationship quality | 0.69 (0.36, 1.31) | 0.253 | 0.68 (0.28, 1.64) | 0.385 | 0.23 (0.14, 0.37) | **0.000** | 0.26 (0.15, 0.48) | **0.000** |
| **VAC related factors** |  |  |  |  |  |  |  |  |
| Father's nurturing care | 3.80 (1.72, 8.39) | **0.001** | 3.95 (1.33, 11.71) | **0.013** | - | - | - | - |
| Caregiver's positive reinforcement | 1.73 (0.83, 3.60) | 0.145 | 2.57 (0.86, 7.65) | 0.090 | - | - | - | - |
| Confidence in avoiding violent discipline toward U5 child | 0.43 (0.26, 0.72) | **0.001** | 0.36 (0.17, 0.77) | **0.008** | - | - | - | - |
| Attitudes toward violent discipline on U5 children | 0.62 (0.29, 1.34) | 0.222 | 0.97 (0.37, 2.57) | 0.948 | - | - | - | - |
| VAC descriptive norms | 0.95 (0.52, 1.74) | 0.868 | 1.40 (0.57, 3.51) | 0.476 | - | - | - | - |
| VAC injunctive norms | 1.59 (0.95, 2.67) | 0.079 | 0.75 (0.30, 1.87) | 0.537 | - | - | - | - |
| **IPV related factors** |  |  |  |  |  |  |  |  |
| Coercive behavior toward wife | - | - | - | - | 7.92 (3.62, 17.34) | **0.000** | 5.23 (2.26, 12.13) | **0.000** |
| Attitudes toward IPV | - | - | - | - | 4.18 (2.61, 6.69) | **0.000** | 2.92 (1.66, 5.14) | **0.000** |
| IPV descriptive norms | - | - | - | - | 1.83 (1.28, 2.63) | **0.001** | 1.55 (1.02, 2.36) | **0.038** |
| IPV injunctive norms | - | - | - | - | 1.51 (1.00, 2.29) | **0.048** | 0.99 (0.60, 1.63) | 0.960 |
| <sup>a</sup> Models adjusted for age, education, HH food insecurity, total children in HH & VAC covariates in table |  |  |  |  |  |  |  |  |
| <sup>b</sup> Models adjusted for age, education, HH food insecurity, total children in HH & IPV covariates in table |  |  |  |  |  |  |  |  |

Among women’s sociodemographic characteristics, higher education, (aOR=0.47; p<0.01) fewer children (aOR=4.41, p<0.01), and higher reported household food insecurity (aOR=0.44, p<0.05) were significantly associated with perpetration of physical VAC in bivariate and multivariate models. Unlike men, attitudes and norms were not significantly associated with physical VAC for women. Only increasing confidence that mothers could avoid violent discipline towards their under-five children (aOR=0.36, p<0.01) was significantly associated with a decreased likelihood of reporting physical VAC among mothers.

### Bivariate and multivariate analysis for husbands’ perpetration and wives’ experience of IPV behaviors

Table 2 and Table 3 include bivariate descriptive and multivariate logistic regression for men’s and women’s reports of perpetrating and experiencing IPV. Among sociodemographic information for men, increasing number of children in a household (aOR=1.53, p<0.05), being in a polygamous marriage (aOR=2.76, p<0.05), and perceptions of lower relationship quality (aOR=0.38, p<0.01) were associated with perpetrating IPV against their partner in both bivariate and adjusted models. Perpetration of a coercive behavior towards their partner (aOR=3.00, p<0.01) and personal attitudes accepting IPV in their relationship (aOR=3.67, p<0.01) were associated with the likelihood of perpetrating IPV against their partner in multivariate models.

Among women, only relationship quality was significantly (aOR=0.26, p<0.01) and being in a polygamous marriage was significantly (aOR=2.86, p=0.05) associated with experiencing IPV after accounting for potential confounders. Looking at other potential predictors, experiencing coercive behaviors (aOR=5.23, p<0.01), personal attitudes toward acceptance of IPV (aOR=2.93, p<0.01), and perceiving that IPV is common in their communities or perceptions of descriptive norms (aOR=1.55, p<0.05) were significantly associated with experiencing IPV in the previous three months.

*Path analysis model for father’s use of physical violent discipline.* The path analysis (Figure 1) explores the relationship between men’s attitudes and perceptions of descriptive and injunctive norms around IPV and VAC, assessment of their relationship quality, any IPV in the relationship as reported by either the male or female partner, a female partner’s report of physical VAC against an under-five child in the previous three months, and their own reports of physical VAC (the outcome). The fit statistics for the path analysis model for men (Figure 1) are presented in Table 4. This model demonstrates close fit through chi-square test, RMSEA, and SRMR measures of model fit and acceptable fit according to CFI and TLI measures [26].

**Figure 1:**
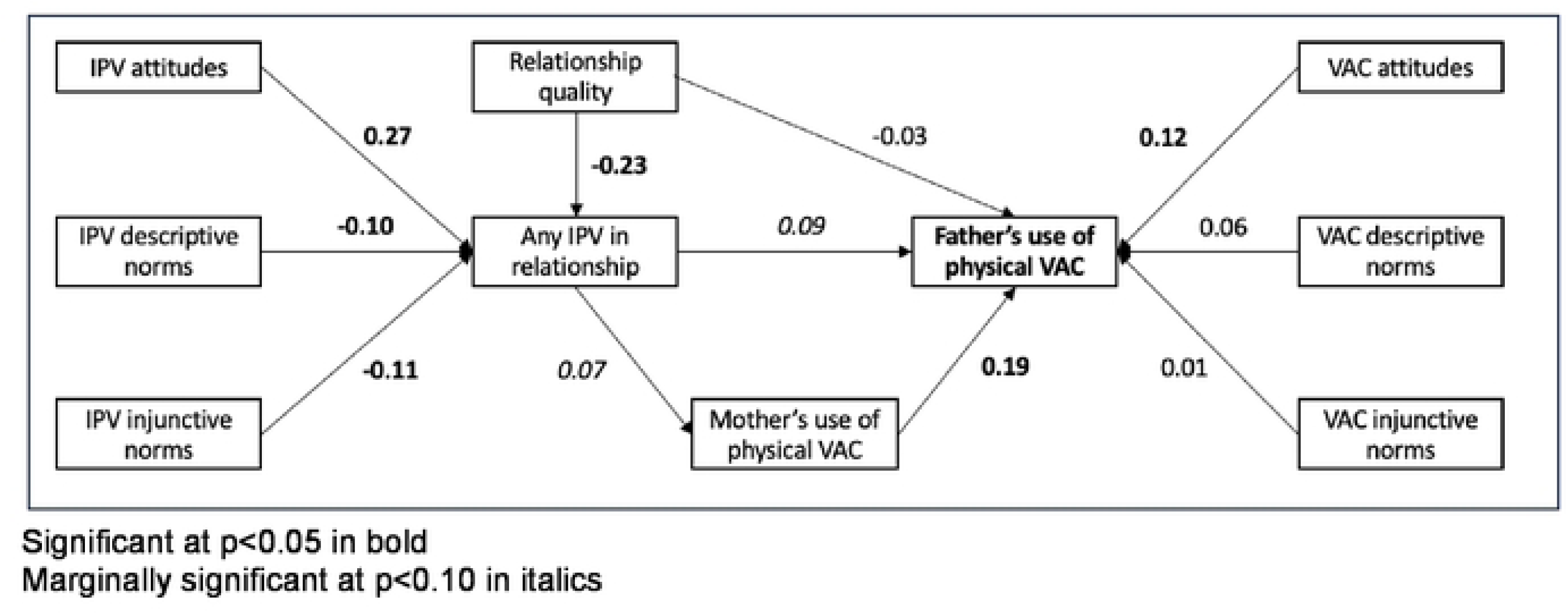
Path model for fathers/husbands; IPV, VAC, attitudes, and norms for father’s use of physical VAC against under-five children in the past three months

**Table 4:** Fit statistics path model for fathers/husbands; IPV, VAC, attitudes, and norms for father’s use of physical VAC against under-five children in the past three months

| Fit statistic | Value for men's SEM model |
| --- | --- |
| *Likelihood ratio* |  |
| p > chi2 | 0.087 |
| *Population error* |  |
| RMSEA | 0.044 |
| *Baseline comparison* |  |
| CFI | 0.861 |
| TLI | 0.743 |
| *Size of residuals* |  |
| SRMR | 0.041 |

We observed a significant (p<0.01) and positive relationship between attitudes supportive of IPV and any reported IPV in the relationship. At the same time, and perhaps a bit unexpectedly, we observed a negative and significant relationship (p<0.05) between descriptive and injunctive norms measures, such that couples with male respondents who perceived that IPV was typical and accepted in their community, were less likely report IPV. Finally, relationship quality was significantly (p<0.01) and inversely associated with IPV occurrence in a relationship.

There were fewer significant relationships looking at attitudes and norms related to VAC. Only VAC attitudes were positively associated (p<0.05) with a father’s report of an act of physical discipline on their under-five children. However, we did see a positive and direct, but marginally significant (p<0.10), pathway between any IPV reported in the relationship and a father’s reported use of physical VAC against their under-five children. In addition, we saw a positive and indirect pathway between any reported IPV in a relationship (p<0.10) and a mother’s reported use of physical VAC against their under-five children and a father’s reported use of physical VAC against their under-five children (p<0.01).

## Discussion

In this study, we looked at young men and women around key transition points in their lives – marriage and childbearing – living in two Communes of Southeastern Senegal, with higher rates of poverty compared to other regions of Senegal [5]. This baseline study took place in 2021, amid the COVID-19 epidemic; a time when families may have experienced disrupted household dynamics, including increased economic and household food insecurity and stress, which could result in higher rates of IPV and VAC [27.28]. Our study is one of the few existing studies in sub-Saharan Africa which applies a dyadic perspective to the study of IPV and VAC rather than focusing solely on individuals of one sex [29]. Caregivers in our study reported high rates of physical VAC – reported by over two-thirds of fathers and 80% of mothers – against under-five children in the previous three months. This is considerably higher compared to other studies done in other regions of Southern Senegal [14, 30], but in line with others using similar measures of VAC asking caregivers to report on a wider range of physically violent acts [15, 31]. We found that mothers were more than twice as likely to report an act of psychologically violent discipline compared to fathers and nearly 20% more likely to report an act of physically violent discipline. Moreover, mothers were more than four times more likely to report an act of harsh physical discipline compared to fathers. Studies examining the sex of the caregiver in assessing VAC are sparse, particularly for caregivers of very young children. Studies that have investigated caregiver sex have generally found that mothers are more likely to report using all types of VAC compared to fathers [15, 18, 30]. Carlson et al. [32] posits that this could be influenced by women being more likely to be charged with day-to-day child rearing and disciplining young children, gendered differences in reporting acts of violence, and the gendered burden of co-occurrence of VAC and IPV.

For fathers, we did find that attitudes and injunctive norms were significantly associated with perpetrating physical VAC in multivariate models. In contrast, women who had higher education, higher household food insecurity, and greater confidence in handling their children without violence were less likely to report physical VAC, and women with more children in the household were more likely to report physical VAC. The finding that higher reported household food insecurity was associated with lower reporting of physical VAC by mothers was surprising given findings from reviews on the association [33]. Most fathers and mothers in this study who report using violent discipline also were likely to report acts of positive reinforcement, nurturing care, and/or nonviolent discipline, suggesting that these are not mutually exclusive and that caregivers often utilize multiple methods of violent and nonviolent discipline [34, 35].

This study had high reports of psychological and physical discipline by both fathers and mothers meaning many children are likely experiencing multiple forms of violence from both parents. We do not know if the intensity or frequency of mothers and fathers’ harsh discipline is similar with their child(ren) or if there are differences by a child’s gender. Violent forms of discipline at a young age increases the risk of acute and long-term poor health, socioemotional, cognitive, and behavioral outcomes for children [36, 37]. Understanding more about the circumstances and for which behaviors parents use different forms of discipline could help identify intervention opportunities that address these behaviors.

Almost one-half of women in this study reported past three-month experience of any act of IPV and more than 80% reported experiencing coercive behaviors. Men reported perpetration of each type of IPV with considerably less frequency – about one-half as often for emotional and physical IPV and about one-quarter as often for sexual IPV – compared to their female partners reporting on their experiences over the same time frame. This study focused on younger women, early in their marital relationship, and confirms data from other studies that IPV is prevalent at young ages and early stages of relationship [2]. Research in other settings found similar risk factors for IPV including being in a polygamous relationship, having more children, having poor relationship quality, and holding attitudes and perceptions of social expectations allowing for IPV and men’s use of coercive behaviors [38, 39]

A study in Uganda examined how social norms contribute to IPV, finding that IPV is normalized within certain boundaries and considered a private domestic matter [40]. As a result, use of violence in particular circumstances may be considered typical, a demonstration of care, and a normal action as head of the family. In our study, descriptive and injunctive norms were associated with perpetrating and experiencing IPV. However, only descriptive norms held by women remained significant after accounting for other factors. We expected strong normative associations with IPV and VAC given the population and assumptions of the REAL Fathers intervention. It is possible that our social norms measures are inadequate given that few reliable and valid normative measures for violence currently exist, especially for sub-Saharan Africa [41].

In households in this setting, violence was a common experience with 71% of men and 89% of women reporting any use of violent discipline in the past three months and 49% of women reporting IPV experience. In other words, in almost all households where women reported IPV, they also used violent discipline, and many women used violence as a form of discipline even without personally experiencing IPV. These findings are supported by the path analysis which focused on husbands/fathers as perpetrators of violence against their partners and under-five children in the previous three months. The path analysis indicated a positive relationship between men’s attitudes allowing for IPV and poor relationship quality with IPV in the relationship. Unexpectedly, where descriptive norms and injunctive norms permitted IPV, IPV was less likely to be reported in the relationship. While the study cannot fully explain this relationship, and it contradicts the multivariate regression, it is possible that norms related to the acceptability of IPV results in under-reporting of IPV, possibly because men and women do not report or note their use or experience of the behavior because of its normativeness. It is also possible that our norms measures are inadequate for this analysis.

The path analysis did show a relationship between IPV, mothers use of physically violent discipline and father’s use of physically violent discipline. Given the commonness of violent discipline by both men and women in this population and the shared beliefs on parenting and discipline as shown by men and women’s attitudes and reported norms, this relationship aligns with expectations. Furthermore, there is widespread evidence to suggest that these multiple forms of violence co-occur within a household, both within and across generations [21, 32, 42]. Reviews from low- and middle-income countries (LMIC) for both types of violence suggest that IPV and violence against children (VAC) share many risk factors such as social norms which condone violence, gender inequality and norms, and economic distress, among others [21, 32, 42]. Currently, there is a small but growing evidence base looking at the co-occurrence of violence in LMIC [43]. Our study contributes to this gap by exploring a range of risk factors which might be associated with co-occurrence of IPV and VAC in LMIC. It provides further support for integrated interventions, working with both fathers and mothers, to prevent violence given the connection between different forms of violence and the multiple factors upholding its use.

This study has some limitations. First, the village sites were purposively sampled and only men who were likely to complete the REAL Fathers intervention were selected for the intervention and study. Therefore, this sample may not be generalizable to all men and women in this setting, and may underestimate the prevalence of IPV and VAC. All questions, including those pertaining to IPV and VAC, were based on self-report and there is a possibility of social desirability bias and under-reporting of household violence, particularly among men. Indeed, we saw large disparities in couples with considerably fewer men reporting perpetrating IPV and VAC compared to women. Among women, it is also possible that violence was under-reported if they feared disclosure of their responses. This disparity persisted in the findings despite our efforts to allow for anonymous responses for IPV and VAC questions and enhanced protections of privacy and data security. This study is also cross-sectional and does not establish causality or directionality in the relationships assessed. While we used validated measures when available, not all of our measures – particularly our social norms measures – have been validated in the Senegal setting. Finally, the lack of qualitative research as part of this evaluation study limited our ability to triangulate and unpack these relationships further.

## Conclusion

Our study contributes to the growing evidence base looking at the intersection of IPV and VAC in LMIC households. It demonstrates high co-occurrence of both forms of violence and finds positive associations between women’s experience of IPV, men’s perpetration of IPV, and both mother’s and father’s use of violence against under-five children in rural Southern Senegalese households. Our study suggests strong influences of personal attitudes and social norms for a young population recently undergoing marriage and childbearing, sometimes in unexpected ways. Ultimately, this study provides support for integrated interventions which simultaneously addresses both IPV and VAC.

## Data Availability

All data are in the manuscript and supporting information files.

## Acknowledgements

The authors would like to thank the men and women who participated in REAL Fathers and this baseline study. We are grateful to the interviewers and supervisors who conducted this study and to local organizations and individuals, Plan Senegal, and government officials who supported this study.

